# Effects of Sacred Dances versus Conventional Physical Training on Functional Capacity and Quality of Life in Women with Heart Failure: Protocol for a Randomized Active-Controlled Crossover Non-Inferiority Trial (RED-WHF)

**DOI:** 10.64898/2026.11.10.26367781

**Authors:** Maria do Rosário Toscano von Flach, Elisabete Silva Fernandes, Céu Barreiros d’Almeida, Humberto Caldas Suarez, Marina von Flach Correia, Ana Amélia Lemos Junquilho, Ana Maria Ferreira das Neves Abreu, Cristiane Maria Carvalho Costa Dias, Luiz Eduardo Fonteles Ritt

## Abstract

This protocol describes the RED-WHF trial, a randomized, active-controlled, two-period crossover clinical trial designed to evaluate whether Sacred Dances (SD)—as a mind-body intervention—are non-inferior to conventional physical training (PT) regarding functional capacity and quality of life in women with heart failure (HF). Sedentary behavior and emotional stress significantly worsen HF prognosis, yet conventional exercise programs face barriers in patient adherence and variable motivation. Sacred Dances, a form of low-to-moderate-intensity moving meditation, may enhance physical fitness while promoting emotional well-being and spiritual connection (flourishing). We will recruit 30 sedentary women aged 18 to 70 with stable HF (NYHA class I-III) and a documented left ventricular ejection fraction. Participants will be randomized 1:1 to one of two treatment sequences: SD followed by PT, or vice versa. Each 12-week intervention phase consists of two weekly sessions, lasting 90 minutes for SD (with 60 minutes of active dancing to allow choreography instruction) and 60 minutes for PT, separated by a 3-week washout period. Primary outcomes are functional capacity (6-minute walk test [6MWT]) and health-related quality of life (EQ-5D-5L Index and VAS). Secondary outcomes include adherence rates, spiritual attitudes (ARES scale), symptoms of depression, anxiety, and stress (DASS-21), and perceived subjective impact (EUPI-12). Statistical planning incorporates linear mixed models (LMM) with treatment, period, and sequence fixed effects, and an intention-to-treat approach with multiple imputation for missing data. To establish non-inferiority, a 30-meter margin for the 6MWT difference will be applied. The study was approved by the local Ethics Committee (CEP/EBMSP opinion 7.443.180, CAAE: 86133125.0.0000.5544) and prospectively registered in the Brazilian Registry of Clinical Trials (ReBEC ID: RBR-85tkdz5).

## Introduction

Cardiovascular diseases (CVDs) remain the leading cause of mortality worldwide, representing a major public health challenge [1, 2]. With the progressive aging of the global population, the prevalence of heart failure (HF) has risen dramatically, affecting millions of individuals [3, 4].

Approximately half of this clinical population consists of women, who frequently suffer from more pronounced functional limitations and worse health-related quality of life (HRQoL) in advanced age compared to men [5, 6].

Sedentary behavior and chronic emotional stress act synergistically to exacerbate the pathophysiological progress of HF, causing endothelial dysfunction, chronic inflammation, and elevated sympathetic drive [6, 7]. Although regular physical exercise is strongly recommended by international guidelines to improve exercise tolerance and reduce hospitalizations [7, 8], conventional cardiac rehabilitation programs face substantial real-world barriers. Most notably, lifestyle modifications and exercise compliance often suffer from low and highly variable motivation among both patients and healthcare providers, leading to elevated drop-out and attrition rates [8, 9].

To overcome these motivational barriers, mind-body interventions (MBIs) have emerged as powerful complementary tools [10, 11]. MBIs are defined as practices that promote interactions between the brain, mind, body, and behavior to enhance physical and mental health [10]. In Brazil, these practices have been officially integrated into the Unified Health System (SUS) since 2006 under the National Policy of Integrative and Complementary Practices (PNPIC) [12]. Cumulative evidence suggests that MBIs can reduce cardiovascular risk factors, improve HRQoL, and decrease adverse cardiac events [13, 14, 15]. In HF populations specifically, MBIs such as Yoga, Tai Chi, and meditation have been shown to reduce physical and psychological symptoms while improving clinical outcomes [15, 16].

Sacred Dances (SD), established by Bernhard Wosien in 1976 in the Findhorn Community, Scotland, represent a unique and understudied form of active moving meditation [16, 17]. Performed mostly in circles, these traditional and contemporary choreographies require no previous physical or artistic training and utilize repetitive movements that promote psychomotor skills, balance, cognition, and psycho-emotional well-being [17, 18]. Furthermore, when integrated with Seligman’s Flourishing framework (PERMA model) [29], Sacred Dances target holistic human development—fostering positive emotions, engagement, relationships, meaning, and accomplishment [29, 30, 31].

This protocol describes the design of the RED-WHF trial, which is the first randomized active-controlled clinical trial to systematically evaluate the functional, clinical, psychological, and spiritual impacts of Sacred Dances in women with heart failure. We hypothesize that Sacred Dances are non-inferior to conventional multicomponent physical training in improving functional capacity and health-related quality of life.

## Methods

### Study Design and Reporting Compliance

The RED-WHF trial is a randomized, active-controlled, two-period, two-sequence crossover clinical trial. The study protocol has been developed in strict compliance with the SPIRIT 2025 statement (Fig 1, S1 Checklist) [36] and its relevant extensions for crossover randomized trials and non-inferiority trial designs. The full institutional trial protocol approved by the Research Ethics Committee is provided as S1 Appendix (English translation) and S2 Appendix (Portuguese original). Detailed intervention guides and session manuals for both arms are provided in S3 Appendix. The study is prospectively registered in the Brazilian Registry of Clinical Trials (ReBEC ID: RBR-85tkdz5; primary registry of the WHO ICTRP network; S4 Appendix).

**Fig 1.**
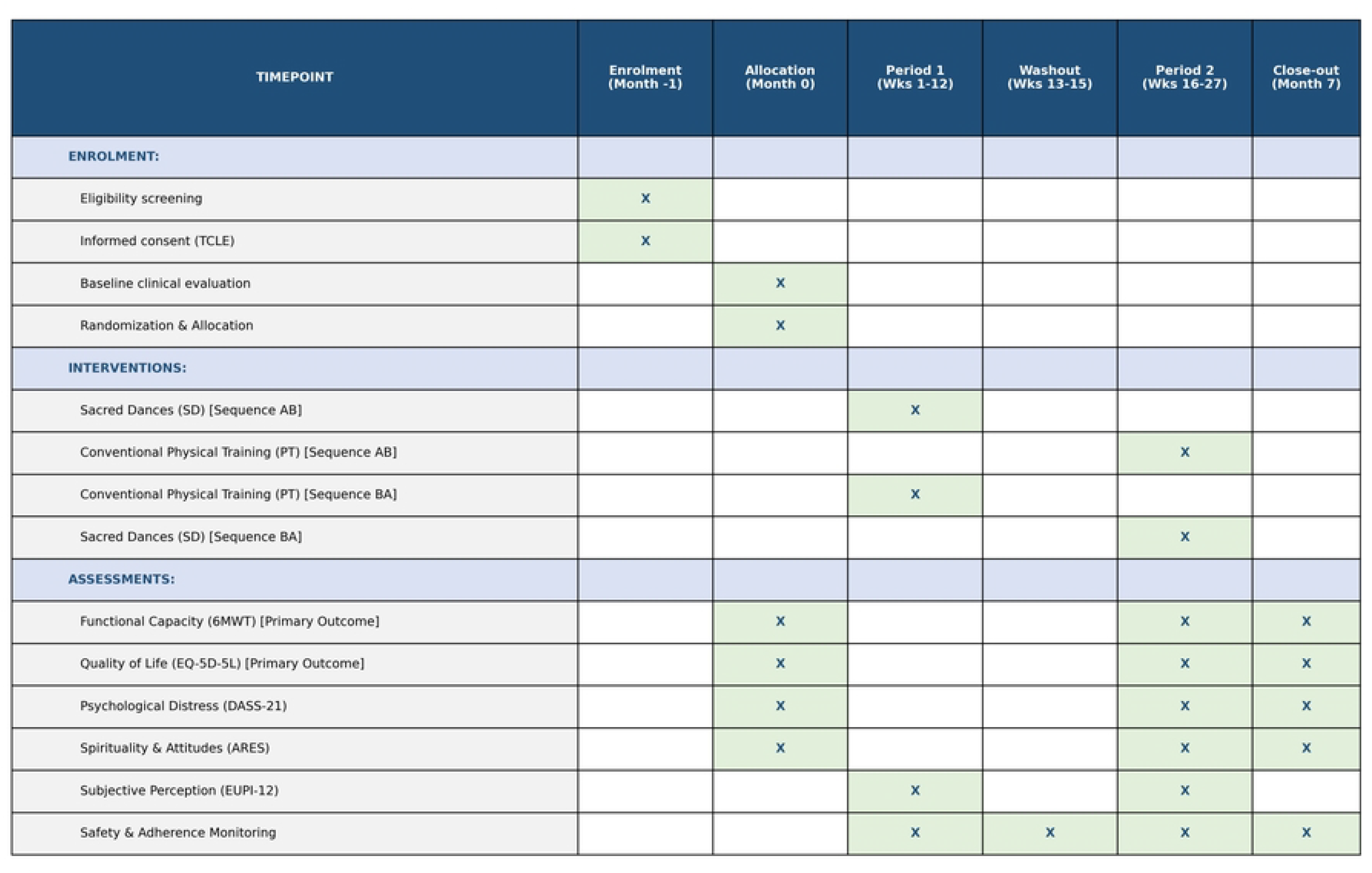
SPIRIT Schedule of Enrolment, Interventions, and Assessments. Schedule of study activities across enrolment, allocation, 12-week intervention periods (Period 1 and Period 2), 3-week washout period, and close-out evaluation for the RED-WHF trial.

### Study Setting and Patient Eligibility

The study will be conducted at the Ambulatório Docente Assistencial of the Escola Bahiana de Medicina e Saúde Pública (EBMSP) in Salvador, Bahia, Brazil. We will recruit sedentary women aged 18 to 70 years with a documented diagnosis of stable chronic heart failure (NYHA functional class I, II, or III) of any etiology and a documented left ventricular ejection fraction (LVEF).

Inclusion criteria comprise: (1) female sex; (2) age between 18 and 70 years; (3) confirmed diagnosis of chronic HF (NYHA class I-III) with stable cardiologist follow-up; (4) sedentary status, defined according to the International Physical Activity Questionnaire (IPAQ) [21] as not having engaged in regular physical exercise for at least 10 continuous minutes per week over the preceding 3 months; and (5) ability to perform the proposed physical activities. Exclusion criteria include: (1) clinical conditions that prohibit physical exercise (e.g., unstable angina, acute myocardial infarction or cardiac surgery within the preceding 8 weeks, complex uncorrected arrhythmias, severe uncompensated valvular disease); (2) severe pulmonary limitations (such as severe uncontrolled asthma or COPD); (3) severe renal failure undergoing hemodialysis; (4) major cognitive or psychiatric impairments that prevent understanding or signing the informed consent; and (5) inability to attend the scheduled group sessions.

### Randomization, Allocation Concealment, and Blinding

Eligible participants who provide written informed consent will be randomized in a 1:1 ratio to one of two intervention sequences: Sequence AB (Sacred Dances in Period 1, followed by a 3-week washout, then Conventional Physical Training in Period 2) or Sequence BA (Conventional Physical Training in Period 1, followed by a 3-week washout, then Sacred Dances in Period 2). Randomization will be stratified by baseline NYHA functional class (Class I-II vs. Class III) to ensure clinical balance across sequences. The random allocation sequence will be generated using a computer-generated random block algorithm embedded in the secure REDCap (Research Electronic Data Capture) platform.

Given the nature of physical exercise and movement interventions, blinding participants and session instructors is impossible. However, rigorous measures will be implemented to prevent assessment bias. An independent, trained health professional (Ana Amélia Lemos Junquilho), completely uninvolved in recruitment, sequence generation, or session delivery, will serve as the blinded outcome assessor. This assessor will remain strictly blinded to sequence allocations throughout the trial and will independently conduct all functional physical tests (6MWT) and administer psychometric instruments (EQ-5D-5L, DASS-21, ARES, EUPI-12). The statistician conducting the final data analysis will also be blinded, working with an alphanumeric-coded database that conceals group identities.

### Study Interventions

Both intervention programs (Sacred Dances and Conventional Physical Training) were structured to ensure physiological equivalence in terms of weekly frequency (2 sessions per week for 12 weeks), overall duration, inclusion of warm-up and cool-down phases, and target metabolic expenditure (light-to-moderate intensity, 2 to 6 METs; Borg CR10 rating of 3-6) [32, 33]. Detailed intervention guides and session manuals are provided in S3 Appendix.

Intervention A (Sacred Dances): Consists of 90-minute sessions held twice weekly for 12 weeks. To ensure metabolic equivalence with the 60-minute exercise circuit, each 90-minute session includes approximately 60 minutes of active dancing, reserving 30 minutes for choreography instruction, demonstration, and group reflection. The 12-session curriculum was co-designed by the PI and certified facilitators (Elisabete Silva Fernandes and Céu Barreiros d’Almeida), combining traditional and contemporary circle choreographies [17, 30, 31]. The energy expenditure for each dance was calculated based on rhythm, tempo, and specific movement patterns (e.g., short steps, arm movements, displacement steps), maintaining an intensity of 2 to 6 METs [32]. To prevent cognitive distraction from native language lyrics, non-Brazilian instrumental or foreign-language tracks are utilized.

Intervention B (Conventional Physical Training): Consists of 60-minute structured multicomponent exercise sessions held twice weekly for 12 weeks, conducted by a certified physical education specialist (Humberto Caldas Suarez and Marina von Flach Correia). Each session comprises: (1) 10 minutes of dynamic warm-up and joint mobility; (2) 40 minutes of a calisthenics and resistance circuit utilizing body weight, elastic resistance bands, agility ladders, and light props (1-3 sets of 10-15 repetitions targeting major muscle groups, interspersed with dynamic aerobic exercises like jumping jacks/march); and (3) 10 minutes of cool-down, static stretching, and diaphragmatic breathing [8, 34, 35]. The intensity is individually monitored using the Borg CR10 scale to maintain effort within 3 to 6 (‘moderate’) while strictly avoiding the Valsalva maneuver.

### Study Variables and Outcomes

The primary outcomes of the trial are: (1) Functional Capacity, evaluated by the total distance walked (in meters) during the standardized 6-minute walk test (6MWT), conducted according to ATS guidelines [22, 23]; and (2) Health-Related Quality of Life (HRQoL), evaluated using the validated Brazilian Portuguese version of the EQ-5D-5L questionnaire [24, 25, 26], including both the 5-dimension descriptive index and the Visual Analogue Scale (VAS, 0–100).

Secondary outcomes include: (1) Psychological Distress, screened using the 21-item Depression, Anxiety and Stress Scale (DASS-21) [27]; (2) Spiritual Attitudes, evaluated using the unidimensional 11-item Attitudes Related to Spirituality Scale (ARES) [28], treated as a continuous covariate and secondary outcome; (3) Intervention Adherence, measured as the percentage of attended sessions over each 12-week period; and (4) Perceived Subjective Impact, evaluated using the study-specific, exploratory 12-item Unified Scale of Perceived Impact of the Intervention (EUPI-12) as a mixed-methods quantitative framework to complement and structure qualitative interview transcriptions.

### Sample Size Calculation and Statistical Planning

The sample size was calculated for the primary outcome of functional capacity (6MWT distance) in a two-period crossover non-inferiority design [19, 20]. Based on cardiac literature, we assumed an expected mean 6MWT distance of 300 meters in both interventions with an inter-individual standard deviation (σ) of 50 meters. Assuming an intra-subject correlation (ρ) of 0.5, the standard deviation of intra-individual differences (σd) is 50 meters. The non-inferiority margin (Δ) was set at 30 meters, representing a clinically accepted minimal important difference threshold for female HF populations [19, 20].

Assuming a one-sided significance level (α) of 2.5% and statistical power (1-β) of 80%, the formula for two-period crossover non-inferiority comparison [n = ((z_1−_α + z_1−_β)^2^ × σd^2^) / Δ^2^] yielded a minimum sample size of 22 participants. To account for a potential 10% dropout rate, the sample size was expanded to 30 participants. Sample size parameters and power estimations were verified using R statistical software (pwr / TrialSize packages).

Statistical analysis will be conducted using linear mixed models (LMM) to account for the repeated-measures crossover structure [36]. Models will include fixed effects for treatment (Sacred Dances vs. Physical Training), period (Period 1 vs. Period 2), and sequence (AB vs. BA), with a random intercept for each participant. Non-inferiority will be established if the lower bound of the one-sided 95% confidence interval (or two-sided 90% CI) for the mean difference in 6MWT distance (SD minus PT) does not exceed -30 meters. If non-inferiority is demonstrated, an exploratory test for superiority will be applied (p < 0.05). All analyses will follow the Intention-to-Treat (ITT) principle, with missing data handled via multiple imputation or linear mixed effect likelihood estimation.

### Ethics and Dissemination

The trial is conducted in accordance with the Declaration of Helsinki and Resolution 466/2012 of the Brazilian National Health Council. The study has been formally approved by the Research Ethics Committee (CEP) of the Escola Bahiana de Medicina e Saúde Pública under opinion number 7.443.180 (CAAE: 86133125.0.0000.5544) on March 14, 2025. The Principal Investigator will personally explain the trial objectives, procedures, risks, and benefits, and obtain written informed consent (TCLE) from each participant on the day of the baseline clinical evaluation.

Participant data will be completely anonymized and securely managed on the REDCap platform. To protect participants, free psychological support will be provided by collaborating psychologists in case of emotional distress, and immediate medical care is guaranteed by the clinical emergency team at the EBMSP facility (Ambulatório Docente Assistencial). Trial findings will be published in peer-reviewed scientific journals, presented at academic congresses, and deposited in open science repositories (OSF).

### Roles and Responsibilities

The Principal Investigator (Maria do Rosário Toscano von Flach) is responsible for overall study design, funding, clinical oversight, ethical submissions, medical screening, safety monitoring, and data management. The Sacred Dance intervention was co-designed and developed with the volunteer facilitators Elisabete Silva Fernandes and Céu Barreiros d’Almeida. The Conventional Physical Training protocol was designed by Humberto Caldas Suarez and Marina von Flach Correia (certified physical education specialists). The independent blinded outcome assessor is Ana Amélia Lemos Junquilho, who is also responsible for critical text review.

Academic advisors Luiz Eduardo Fonteles Ritt, Cristiane Maria Carvalho Costa Dias, and Ana Maria Ferreira das Neves Abreu supervise methodological rigor, clinical safety, and ethical compliance across all phases.

### Declaration of Generative AI and AI-Assisted Technologies

During the preparation of this manuscript, the authors used generative AI tools (specifically OpenEvidence and Gemini Notebook) exclusively for searching scientific literature, verifying calculations, adjusting formatting, and refining language translation. Following the use of these tools, all content was thoroughly reviewed, edited, and verified by the authors, who assume full responsibility for the final contents of the published manuscript.

## Data Availability

All data produced in the present study are available upon reasonable request to the authors

https://github.com/emiliehuon/uterus_analysis_contractility,

## Supporting Information

S1 Appendix. Full Original Trial Protocol (English Translation). Complete clinical trial protocol approved by the Research Ethics Committee (CEP/EBMSP Opinion No. 7.443.180 and Amendment No. 8.405.010/E1), translated into English.

S2 Appendix. Full Original Trial Protocol (Portuguese Original). Complete clinical trial protocol approved by the Research Ethics Committee (CEP/EBMSP Opinion No. 7.443.180 and Amendment No. 8.405.010/E1) in Portuguese.

S3 Appendix. Detailed Intervention Guides and Session Manuals. Step-by-step practical session guides and instructional manuals for Sacred Dances and Conventional Physical Training.

S4 Appendix. Clinical Trial Registration. Confirmation of primary clinical trial registration in the Brazilian Registry of Clinical Trials (ReBEC ID: RBR-85tkdz5; WHO ICTRP network).

S1 Checklist. SPIRIT 2025 Checklist. Completed SPIRIT 2025 checklist for clinical trial protocol reporting.

